# Explore the Possible Impact of BCG Vaccination Policy on the Morbidity, Mortality, and Recovery Rates due to COVID-19 Infection

**DOI:** 10.1101/2020.06.14.20131268

**Authors:** Chuan-Hsin Chang, Yue-Cune Chang

## Abstract

**BACKGROUND:** The Coronavirus Disease-19 (COVID-19) is the new form of an acute infectious respiratory disease and has quickly spread over most continents in the world. Recently, it has been shown that Bacille Calmette-Guerin (BCG) might protect against COVID-19. This study aims to investigate the possible correlation between BCG vaccination and morbidity/mortality/recovery rate associated with COVID-19 infection.

**METHODS:** Data of COVID-19 confirmed cases, deaths, recoveries, and population were obtained from https://www.worldometers.info/coronavirus/ (Accessed on 12 June, 2020). To have meaningful comparisons among countries’ mortality and recovery rates, we only choose those countries with COVID-19 infected cases at least 200. The Poisson regression and logistic regression were used to explore the relationship between BCG vaccination and morbidity, mortality and recovery rates.

**RESULTS:** Among those 158 countries with at least 200 COVID-19 infected cases, there were 141 countries with BCG vaccination information available. The adjusted rates ratio of COVID-19 confirmed cases for Current BCG vaccination vs. non-Current BCG vaccination was 0.339 (with 95% CI= (0.338,0.340)). Moreover, the adjusted odds ratio (OR) of death and recovery after coronavirus infected for Current BCG vaccination vs. non-Current BCG vaccination were 0.258 (with 95% CI= (0.254,0.261)) and 2.151 (with 95% CI= (2.140,2.163)), respectively.

**CONCLUSIONS:** That data in this study show the BCG might provide the protection against COVID-19, with consequent less COVID-19 infection and deaths and more rapid recovery. BCG vaccine might bridge the gap before the disease-specific vaccine is developed, but this hypothesis needs to be further tested in rigorous randomized clinical trials.

## Introduction

The Coronavirus Disease-19 (COVID-19) pandemic is caused by severe acute respiratory syndrome coronavirus 2 (SARS-COV-2), which is a single-stranded positive-sense RNA virus. The outbreak was first identified in Wuhan, China, and it has quickly spread over all continents in the world. At the time of writing, there are more than 427 thousand people die of COVID-19 and more than 7.7 million people infected. The number of cases is still rising without any sign of stopping.

Recently, there are several studies mention Bacille Calmette-Guerin (BCG) might protect against COVID-19.^1,2,3^ It has been shown that the earlier that the countries established the BCG vaccination policy, the stronger reduction in the numbers of the deaths due to COVID-19 infection.^1^ Also, BCG vaccination slowed down the spread or progression of symptoms and death due to COVID-19.^2,3^

BCG is a live attenuated vaccine derived from a strain of *Mycobacterium bovis* primarily used against tuberculosis (TB). Many nations, including Taiwan, Japan, and China, have a universal BCG vaccination policy in newborns. While other countries such as Spain, France, and Switzerland discontinued universal BCG policies. The US, Italy and the Netherlands have not adopted universal BCG requirements.

In addition to the specific effect of BCG vaccination against TB, BCG has the beneficial non-specific (heterplogous) effects on the immune system and induces cross-protection against other mycobacterial pathogens. The studies show that BCG reduced the incidence of acute lower respiratory tract infection (ALRI) and pneumonia,^4–6^ sepsis- and pneumonia-related neonatal mortality,^7^ yellow fever vaccine viremia,^8^ as well as was used for the treatment of bladder cancer.^9,10^ In animal model, BCG reduced viral titers of influenza A virus (H7N9)^11^ and protected from herpes simplex virus type 2 (HSV2).^12^ And BCG markedly reduced the severity of mengovirus (encephalomyocarditis virus) infection in mice.^13,14^ The potential cellular and molecular mechanism of non-specific effect of BCG against viral infection has been studies only in the last decades. BCG vaccination significantly increases the secretion of pro-inflammatory cytokines such as IL-1β, tumor necrosis factor (TNF), IFN-γ and IL-6.^15,16^ That might be accompanied with the transcriptional, epigenetic and metabolic reprogramming of innate immune cells, and the phenotypic change in the innate immune cell induces innate immune memory called “trained immunity”.^8,17,18^ Therefore, the “trained immunity” induced by BCG vaccination may have a role in protecting against COVID-19 virus.

This study aims to investigate the possible correlation between BCG vaccination and morbidity/mortality/recovery rate associated with COVID-19 infection. The development of the effective vaccine might curb the spread of the virus, but that is expected to take at least 12-18 months to develop. Therefore, the cross-protection induced by BCG vaccine might be a bridge to the specific COVID-19 vaccine.

## Method

The definition of BCG vaccination is according to BCG World Atlas 2nd Edition.^19^ Data of COVID-19 confirmed cases, deaths, recoveries, and population were obtained from https://www.worldometers.info/coronavirus/ (Accessed on 12 June, 2020). However, it is understandable that the current number of COVID-19 cases is highly underestimated worldwide due to lack of comprehensive screening especially for those lower income countries, for example India. To be able to adjusted for the effect of lower income countries, the income classification (1: Low income; 2: Lower middle income; 3: Upper middle income; 4: High income) was based on World Bank list of economies (https://datahelpdesk.worldbank.org/knowledgebase/articles/906519-world-bank-country-and-lending-groups, Accessed on 12 June, 2020). The generalized linear models (GLM) were used to explore the possible factors’ effects on the morbidity, mortality, and recovery rates due to COVID-19 infection. More specifically, the Poisson regression models were used to explore the possible factors, mainly BCG vaccination, on the morbidity rates after adjusting for the effects of populations size (using logarithm of populations as offset) and other possible confounding variables. The logistic regression models were used to explore the possible factors, mainly BCG vaccination, on the mortality and recovery rates among those COVID-19 infected cases after adjusting for the aforementioned confounding variables. To have a meaningful comparison, we only chose those countries with COVID-19 infected cases at least 200. All analyses were done by using the SPSS v26.0 software (SPSS Inc., Chicago, IL, USA). A *p-*value < 0.05 was regarded as statistically significant.

## Results

There were 158 countries with COVID-19 infected cases at least 200. Among them, there were 17 countries without BCG vaccination information available. The total recovered information is unavailable for UK, Spain, Sweden and Netherlands. There were two countries, Uganda and Vietnam, the total deaths were not reported. To investigate whether the Current BCG vaccination countries have lower number of COVID-19 confirmed cases than non-Current BCG vaccination countries, the Poisson regression models were used with population size (in log-scale) as the offset to take into account the possible impact from various population size. As shown in the model 1 of Table 1, after adjusting for the effect of population size, the (intensity) rates ratio (RR) of COVID-19 confirmed cases for Current BCG vaccination vs. non-Current BCG vaccination was 0.135 (with 95% CI= (0.135,0.136)). In other words, the (intensity) rate of COVID-19 confirmed cases for those countries with Current BCG vaccination was 86.5% (=1-0.135) significantly lower than those non-Current BCG vaccination countries (p-value < 0.001), after adjusting for the effect of population size. The number of confirmed cases might be influenced by multiple factors. It has been reported the number of COVID-19 reported cases might be dramatically underestimated around the world due to the lowest rate of diagnostic tests and poor testing quality in lower income countries^20,21^. Accordingly, in order to account for that, we further analyzed with adjustment of the effects of countries’ economies by adding it into the previous Poisson regression model. As shown in the Model 2 of Table 1, the (intensity) rates ratio of COVID-19 confirmed cases for Current BCG vaccination vs. non-Current BCG vaccination was 0.339 (95% CI= (0.338,0.340)). In other words, the (intensity) rate of COVID-19 confirmed cases for those countries with Current BCG vaccination was 66.1% significantly lower than those non-Current BCG vaccination countries (p-value < 0.001), after adjusting for the additional effects of countries’ economic statuses.

**Table 1.**
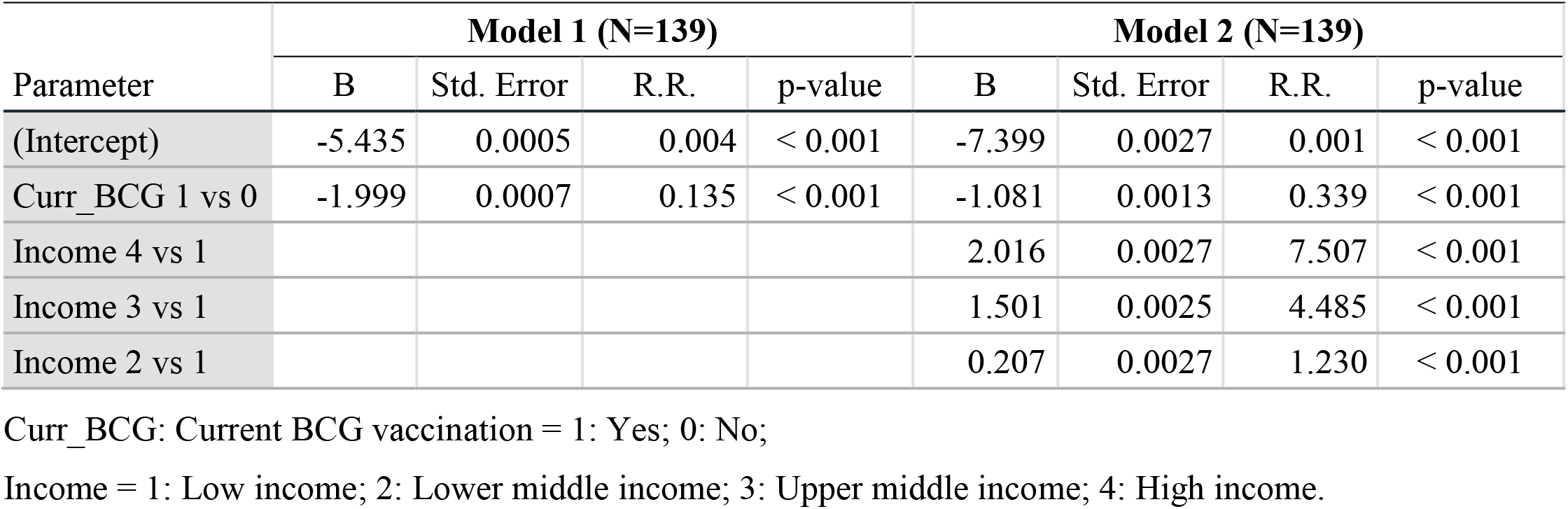
Results of Poisson regression for comparing morbidity rates after adjusting for the population size effect

| Parameter | Model 1 (N=139) |  |  |  | Model 2 (N=139) |  |  |  |
| --- | --- | --- | --- | --- | --- | --- | --- | --- |
|  | B | Std. Error | R.R. | p-value | B | Std. Error | R.R. | p-value |
| (Intercept) | -5.435 | 0.0005 | 0.004 | < 0.001 | -7.399 | 0.0027 | 0.001 | < 0.001 |
| Curr_BCG 1 vs 0 | -1.999 | 0.0007 | 0.135 | < 0.001 | -1.081 | 0.0013 | 0.339 | < 0.001 |
| Income 4 vs 1 |  |  |  |  | 2.016 | 0.0027 | 7.507 | < 0.001 |
| Income 3 vs 1 |  |  |  |  | 1.501 | 0.0025 | 4.485 | < 0.001 |
| Income 2 vs 1 |  |  |  |  | 0.207 | 0.0027 | 1.230 | < 0.001 |

Curr\_BCG: Current BCG vaccination = 1: Yes; 0: No;
Income = 1: Low income; 2: Lower middle income; 3: Upper middle income; 4: High income.

We further used logistic regression to analyze whether BCG vaccination reduced mortality rates caused by COVID-19. Among those (coronavirus) infected (COVID-19 confirmed) cases, the odds ratio (OR) of death for Current BCG vaccination vs. non-Current BCG vaccination was 0.396 (with 95% CI = (0.394,0.399)), as shown in Model 1 of Table 2. In other words, the odds of death due to coronavirus infected for those countries with Current BCG vaccination was 60.4% (=1-0.396) significantly lower than those non-Current BCG vaccination countries (p-value < 0.001), ignoring other factors’ effects. Similarly, as shown in the Model 2 of Table 2, after adjusting for the effects of countries’ economies, the OR of death after (coronavirus) infected for Current BCG vaccination vs. non-Current BCG vaccination was 0.258 (with 95% CI = (0.254,0.261)). In other words, the odds of death due to coronavirus infected for those countries with Current BCG vaccination was 74.2% significantly lower than those non-Current BCG vaccination countries (p-value < 0.001). Accordingly, the data suggests that BCG vaccination seem to significantly reduce mortality associated with COVID-19.

**Table 2.** Results of logistic regression for comparing mortality rates from COVID-19 infected

| Parameter | Model 1 (N=137) |  |  |  | Model 2 (N=137) |  |  |  |
| --- | --- | --- | --- | --- | --- | --- | --- | --- |
|  | B | Std. Error | OR | p-value | B | Std. Error | OR | p-value |
| (Intercept) | -2.447 | 0.0019 | 0.087 | < 0.001 | -2.421 | 0.0107 | 0.089 | < 0.001 |
| Curr_BCG 1 vs 0 | -0.925 | 0.0034 | 0.396 | < 0.001 | -1.356 | 0.0072 | 0.258 | < 0.001 |
| Income 4 vs 1 |  |  |  |  | -0.036 | 0.0109 | 0.965 | < 0.001 |
| Income 3 vs 1 |  |  |  |  | 0.556 | 0.0125 | 1.744 | < 0.001 |
| Income 2 vs 1 |  |  |  |  | 0.200 | 0.0141 | 1.222 | < 0.001 |

Curr\_BCG: Current BCG vaccination = 1: Yes; 0: No;
Income = 1: Low income; 2: Lower middle income; 3: Upper middle income; 4: High income.

We further investigated whether BCG vaccination increases the recovery rate of COVID-19. Among those (coronavirus) infected (COVID-19 confirmed) cases, the OR of recovery for Current BCG vaccination vs. non-Current BCG vaccination was 1.337 (with 95% CI = (1.333,1.341)), as shown in the Model 1 of Table 3. In other words, the odds of recovery from COVID-19 confirmed for those countries with current BCG vaccination was 33.7% (=1.337-1) significantly higher than those non-Current BCG vaccination countries (p-value < 0.001), ignoring other factors’ effects. Similarly, as shown in the Model 2 of Table 3, after adjusting for the effects of countries’ economies, the OR of recovery from coronavirus infection for Current BCG vaccination vs. non-Current BCG vaccination was 2.151 (with 95% CI = (2.140,2.163)). In other words, the odds of recovery from coronavirus infection for those countries with current BCG vaccination was 2.151 times significantly higher than those non-Current BCG vaccination countries (p-value < 0.001), after adjusting for the effects of countries’ economies.

**Table 3.** Results of logistic regression for comparing recovery rates from COVID-19 infection

| Parameter | Model 1 (N=137) |  |  |  | Model 2 (N=137) |  |  |  |
| --- | --- | --- | --- | --- | --- | --- | --- | --- |
|  | B | Std. Error | OR | p-value | B | Std. Error | OR | p-value |
| (Intercept) | -0.060 | 0.0012 | 0.942 | < 0.001 | -0.432 | 0.0051 | 0.649 | < 0.001 |
| Curr_BCG 1 vs 0 | 0.290 | 0.0015 | 1.337 | < 0.001 | 0.766 | 0.0027 | 2.151 | < 0.001 |
| Income 4 vs 1 |  |  |  |  | 0.391 | 0.0052 | 1.478 | < 0.001 |
| Income 3 vs 1 |  |  |  |  | -0.033 | 0.0054 | 0.967 | < 0.001 |
| Income 2 vs 1 |  |  |  |  | -0.739 | 0.0058 | 0.478 | < 0.001 |

Curr\_BCG: Current BCG vaccination = 1: Yes; 0: No;
Income = 1: Low income; 2: Lower middle income; 3: Upper middle income; 4: High income.

## Discussion

Our data have provided the epidemiological evidence indicating that the BCG vaccination might protect against COVID-19. The previous study has shown that the country-by-country difference in COVID-19 morbidity and mortality might be partially explained by national policies on BCG vaccination.^1^ Also, Shet et al. mentioned that the use of BCG might decrease the COVID-19-attributable mortality with adjusting for country economic status and proportion of elderly among the population.^3^ In this study, we adjusted the effect of countries’ economies and population size, there are lower rates of COVID-19 confirmed cases (Table 1), COVID-19 mortality (Table 2), and higher recovery rate of COVID-19 (Table 3) in current BCG vaccination countries compared with non-BCG vaccination countries. However, Sala et al. mentions BCG vaccination has the larger effect on slowing the spread or reducing progression of symptoms than induction of protection from COVID-19 death.^2^ BCG vaccination has been shown to produce broad protection against viral infection and sepsis.^11^ In this study, the data show BCG vaccination caused the reduction in the number of COVID-19 reported infections in the country suggesting BCG might have a specifically protective effect against COVID-19. And the broad use of BCG vaccination across a population could reduce the number of carriers and might slow/stop the spread of COVID-19.

On the basis of these data, we suggest that BCG vaccination might be a potent protective effect on reduction of COVID-19 disease severity. However, there are several confounding factors we might need to further investigate. First, the demographic and genetic structure of the population and the disease burden in different locations; second, the prevention interventions such as social distancing, quarantining, and self-isolating in different countries; third, the difference in report and testing rate of the COVID-19 confirmed cases; forth, the stages of the COVID-19 pandemic in different country; fifth, how long does the induction of “trained immunity” after BCG vaccination last? Sixth, Miyasaka mentioned that different BCG strains may be variably associated with mortality of COVID-19.^22^ The study results might be affected by the different BCG vaccination schedule,^23^ as well as different strains of the bacteria.^24^

Recently, some randomized controlled trials are underway.^25^ In Netherlands, Australia, South Africa, France, and USA, the trials are designed to explore whether BCD-Danish reduces the incidence and severity of COVID-19 in Health Care Workers (NCT04327206, NCT04328441, NCT04379336, NCT04384549, NCT04348370) (see <u>LINK</u>). Also, the clinical trial use BCG as the therapeutic vaccine for COVID-19 confirmed cases in Brazil (NCT04369794). And the observational, case-control, study in Egypt is examining the severity of disease in COVID-19 positive patients who are tested positive for past BCG exposure compared with tested negative (NCT04347876). In summary, the data in this study might support the heterologous immune effects of BCG on prevention of COVID-19 virus infection as well as reduction of morbidity and mortality of COVID-19. The protective effect of BCG vaccination on COVID-19 may be caused by boosting of the innate immune response by BCG. It has been shown that the exaggerated cytokine responses might be associated with complications in patients with COVID-19.^26^ As cautioned by the WHO^27^ and the global shortage of BCG supply,^28^ it might need to further discuss the therapeutic strategy of BCG vaccination. That data in this study show the BCG might provide the protection against COVID-19, BCG vaccine induces trained immunity and provides non-specific protection to bridge the gap before the disease-specific vaccine is developed, but this hypothesis needs to be further tested in rigorous randomized clinical trials.

## Data Availability

The authors confirm that the data supporting the findings of this study are available within the article. These data were derived from the following resources available in the public domains.

https://www.worldometers.info/coronavirus/

https://datahelpdesk.worldbank.org/knowledgebase/articles/906519-world-bank-country-and-lending-groups

## Author Affiliations

From Academic Sinica, Taipei (C.H.C.) and Tamkang University, New Taipei City (Y.C.C)-both in Taiwan.

Address reprint requests to Dr. Yue-Cune Chang at Department of Mathematics, Tamkang University, No.151, Yingzhuan Rd., Tamsui Dist., New Taipei City 25137, Taiwan (R.O.C.), or at

## Reference

1. Miller A, Reandelar MJ, Fasciglione K, Roumenova V, Li Y, Otazu GH. Correlation between universal BCG vaccination policy and reduced morbidity and mortality for COVID-19: an epidemiological study. 2020:2020.03.24.20042937.

2. Sala G, Chakraborti R, Ota A, Miyakawa T. Association of BCG vaccination policy and tuberculosis burden with incidence and mortality of COVID-19. 2020:2020.03.30.20048165.

3. Shet A, Ray D, Malavige N, Santosham M, Bar-Zeev N. Differential COVID-19-attributable mortality and BCG vaccine use in countries. 2020:2020.04.01.20049478.

4. Stensballe LG, Nante E, Jensen IP, et al. Acute lower respiratory tract infections and respiratory syncytial virus in infants in Guinea-Bissau: a beneficial effect of BCG vaccination for girls community based case-control study. Vaccine 2005;23:1251–7.

5. Wardhana, Datau EA, Sultana A, Mandang VV, Jim E. The efficacy of Bacillus Calmette-Guerin vaccinations for the prevention of acute upper respiratory tract infection in the elderly. Acta medica Indonesiana 2011;43:185–90.

6. Nemes E, Geldenhuys H, Rozot V, et al. Prevention of M. tuberculosis Infection with H4:IC31 Vaccine or BCG Revaccination. The New England journal of medicine 2018;379:138–49.

7. Biering-Sørensen S, Aaby P, Lund N, et al. Early BCG-Denmark and Neonatal Mortality Among Infants Weighing <2500 g: A Randomized Controlled Trial. Clinical infectious diseases : an official publication of the Infectious Diseases Society of America 2017;65:1183–90.

8. Arts RJW, Moorlag S, Novakovic B, et al. BCG Vaccination Protects against Experimental Viral Infection in Humans through the Induction of Cytokines Associated with Trained Immunity. Cell host & microbe 2018;23:89-100.e5.

9. Goodridge HS, Ahmed SS, Curtis N, et al. Harnessing the beneficial heterologous effects of vaccination. Nature reviews Immunology 2016;16:392–400.

10. Kamat AM, Porten S. Myths and mysteries surrounding bacillus Calmette-Guérin therapy for bladder cancer. European urology 2014;65:267–9.

11. Moorlag S, Arts RJW, van Crevel R, Netea MG. Non-specific effects of BCG vaccine on viral infections. Clinical microbiology and infection : the official publication of the European Society of Clinical Microbiology and Infectious Diseases 2019;25:1473–8.

12. Starr SE, Visintine AM, Tomeh MO, Nahmias AJ. Effects of immunostimulants on resistance of newborn mice to herpes simplex type 2 infection. Proceedings of the Society for Experimental Biology and Medicine Society for Experimental Biology and Medicine (New York, NY) 1976;152:57–60.

13. Old LJ, Benacerraf B, Clarke DA, Carswell EA, Stockert E. The role of the reticuloendothelial system in the host reaction to neoplasia. Cancer research 1961;21:1281–300.

14. Floc’h F, Werner GH. Increased resistance to virus infections of mice inoculated with BCG (Bacillus calmette-guérin). Annales d’immunologie 1976;127:173–86.

15. Kleinnijenhuis J, Quintin J, Preijers F, et al. Bacille Calmette-Guerin induces NOD2-dependent nonspecific protection from reinfection via epigenetic reprogramming of monocytes. Proceedings of the National Academy of Sciences of the United States of America 2012;109:17537–42.

16. Kleinnijenhuis J, Quintin J, Preijers F, et al. Long-lasting effects of BCG vaccination on both heterologous Th1/Th17 responses and innate trained immunity. Journal of innate immunity 2014;6:152–8.

17. Netea MG, Joosten LA, Latz E, et al. Trained immunity: A program of innate immune memory in health and disease. Science (New York, NY) 2016;352:aaf1098.

18. O’Neill Laj, Netea MG. BCG-induced trained immunity: can it offer protection against COVID-19? Nature reviews Immunology 2020:1–3.

19. BCG World Atlas 2nd Edition. http://wwwbcgatlasorg/indexphp 2017.

20. Curtis N, Sparrow A, Ghebreyesus TA, Netea MG. Considering BCG vaccination to reduce the impact of COVID-19. Lancet (London, England) 2020;395:1545–6.

21. A FLHSMES. Universal BCG vaccination and protection against COVID-19: critique of an ecological study. shttps://naturemicrobiologycommunitynaturecom/users/36050-emily-maclean/posts/64892-universal-bcg-vaccination-and-protection-against-covid-19-critique-of-an-ecological-study 2020.

22. Miyasaka M. Is BCG vaccination causally related to reduced COVID-19 mortality? EMBO molecular medicine 2020.

23. Zwerling A, Behr MA, Verma A, Brewer TF, Menzies D, Pai M. The BCG World Atlas: a database of global BCG vaccination policies and practices. PLoS medicine 2011;8:e1001012.

24. Horwitz MA, Harth G, Dillon BJ, Maslesa-Galić S. Commonly administered BCG strains including an evolutionarily early strain and evolutionarily late strains of disparate genealogy induce comparable protective immunity against tuberculosis. Vaccine 2009;27:441–5.

25. Lawton G. Trials of BCG vaccine will test for covid-19 protection. New scientist (1971) 2020;246:9.

26. Mehta P, McAuley DF, Brown M, Sanchez E, Tattersall RS, Manson JJ. COVID-19: consider cytokine storm syndromes and immunosuppression. Lancet (London, England) 2020;395:1033–4.

27. WHO. Bacille Calmette-Guérin (BCG) vaccination and COVID-19. https://wwwwhoint/news-room/commentaries/detail/bacille-calmette-gu%C3%A9rin-(bcg)-vaccination-and-covid-19 2020.

28. Kuroda N. Demand for BCG Vaccine Due to Unproven Claims of its Role in Preventing COVID-19 Is Causing Shortages of Vaccines for Infants in Japan. The Pediatric infectious disease journal 2020.

